# Effect of Renin-Angiotensin-Aldosterone System inhibitors on outcomes of COVID-19 patients with hypertension: Systematic review and Meta-analysis

**DOI:** 10.1101/2020.09.03.20187393

**Authors:** Tamirat Bekele Beressa, Tamiru Sahilu, Serawit Deyno

## Abstract

**Objective:** This research aimed to systematically review and summarize the influence of Renin-Angiotensin-Aldosterone System (RAAS) inhibitors on the outcome of COVID_19 patients with hypertension.

**Methods:** Electronic databases; PubMed/Medline, CINAHL, the Cochrane Central Register of Controlled Trials, clinical trial.gov, and Google Scholar were searched from 2019 to June 1, 2020. Additionally, the references of identified articles were also searched.

**Results:** A total of 9 articles comprising 3,823 patients were incorporated; 1416 patients on RAAS inhibitors and 3469 on non-RAAS inhibitors. The study demonstrated that the taking of RAAS inhibitors in COVID_19 patients with hypertension significantly reduced mortality where patients on RAAS inhibitors had a 27% decrease of mortality (RR = 0.73 [95% CI: 0.63- 0.85, p< 0.0001, I^2^ = 0%, random-effects model]) compared to those not taking ACEI/ARB. No significant association were observed in disease severity (RR = 0.92 (95% CI: 0.74- 1.14) and hospitalization (WMD = –2.33[95% CI: –5.60, 0.75]), random-effects model.

**Conclusion:** This study supports RAAS inhibitors’ safe use among COVID_19 patients with hypertension.

## Background

Corona viral diseases 2019 (COVID_19) is a pandemic disease originated from Wuhan city of, China from December 2019^1^. The pandemic has continued to debilitate global health and the economy. As of June 7, 2020; 7,005,822 cases of COVID_19 were reported, including 402,678 deaths worldwide^2^. Severe COVID_19 illnesses have consistently been observed in patients with Comorbid illnesses such as cardiovascular and diabetes mellitus^3^. According to the report of the Chinese CDC as of February 11, 2020, 10.5% of deaths occurred in COVID_19 patients with cardiovascular diseases and, 6% of them were patients with hypertension^3^.

Most patients with cardiovascular disease use RAAS inhibitors; commonly angiotensin-converting enzyme inhibitor (ACEI) or angiotensin receptor blocker (ARB)^4^. They are a drug of choice for heart failure, hypertension, chronic kidney disease and, myocardial infraction^5, 6^. Besides inhibition of the formation of angiotensin II; RAAS inhibitors increase the expression of angiotensin-converting enzyme 2 (ACE2); a membrane-bound aminopeptidase that is expressed abundantly in the lungs and the heart^7^. ACE2 plays an important role in the breakdown of angiotensin II (a potent vasoconstrictor) to angiotensin (1–7) (a vasodilator)^8^ which helpful in cardiovascular patients.

Researches reported that severe acute respiratory syndrome coronavirus 2 (SARS-CoV-2) attacks the humans cells through ACE2^9, 10^, and reduces ACE2 expression, resulting imbalance of ACE/Ang II/AT1R axis and ACE2/Ang (1–7)/Mas receptor^11, 12^. Therefore; there are concerns about the use of RAAS inhibitors as a result of an increase in expression of ACE2; a target molecule for SARS CoV-2^13–15^. This led to the hypothesis that the use of RAAS inhibitors might be harmful in patient with COVID_19 especially in patients with comorbidity like hypertension. There are retrospective studies conducted to test this hypothesis and one meta-analysis (preprint)summarized the studies in all patients taking RAAS inhibitors^16^. The meta-analysis included 1842 COVID_19 patients and exhibited a significant 43 % decrease in mortality on the side of patients taking ACEI/ARB^16^. After the meta-analysis several studies were conducted which requires updating the evidence and also there is a need for evidence on hypertensive patients. This study aimed to systematically synthesis the evidence on the effect of RAAS inhibitors on the outcome of COVID_19 patients with hypertension.

## Methods

### Study design

This is a systematic review and meta-analysis conducted using electronic database searches. The review conducted according to the preferred reporting items for Systematic Review and Meta-Analysis Protocol (PRISMA)^17^. The protocol was registered in the (Prospero) International Prospective Register of a systematic review with registration number CDR42020186477.

### Search strategy

The databases such as PubMed, the Cochrane Central Register of controlled trials, clinical http://trial.gov, CINAHIL, and Google scholar were searched from 2019 to June 1, 2020. Additionally, the references of identified articles were also searched. No language limitation was employed. Flow diagram was used to summarize the number of studies identified, screened, excluded, and finally included in the study. Search terms for PubMed were attached under the supplementary document (Appendix 1).

### Study selection

Two reviewers (TB and TS) independently carried out searching for literature and identified relevant studies and sequentially screened their titles and abstracts for eligibility. The full texts of eligible studies were retrieved. Differences in the inclusion of articles were fixed on discussion with the third author (SD). A screening guide was used to ensure that all review authors applied the selection criteria.

### Inclusion criteria

Randomized controlled trials (RCTs) including cluster RCTs, observational studies, and prospective, retrospective comparative cohort studies and case-control studies were considered to be included in the systematic review and meta-analysis. The review considered all patients who were taking ACEI or ARBs alone or in combination for hypertension with COVID-19 patients.

### Exclusion criteria

*In vitro* studies, studies not related to ARB or ACEI for COVID-19 patients with hypertension, studies without extractable data, outcome (death), or severity or hospitalization not clearly reported were excluded from the review.

### Methodological quality assessment

Selected papers were evaluated by two reviewers (TB & TS) for methodological validity before inclusion in the review with the Newcastle-Ottawa quality assessment score (NOS) method. Any differences that arose from the reviewers were fixed through discussion, with a third reviewer (SD).

### Data extraction

Two reviewers (TB and TS) extracted data from the studies using a pre-designed format. Data extracted include first author, the region of study, included population, study design, number of study participants, comparator group, patient status, severity, age (mean, median), gender, interventions, and patients outcomes (number of cases).

### Data analysis and statistical methods

Data analysis was performed by RevMan 5.4 (Copenhagen: the Cochrane Collaboration, 2020) and Stata version 13 (StataCorp, 2013). Risk ratios were used for comparison and reported as 95% confidence interval. P-value ≤ 0.05 was considered statistically significant. The Q-statistic test was used to assess the heterogeneity of the included study and I^2^ statistic was used to indicate the percentage of variation in the studies as a result of heterogeneity instead of chance.

## Results

A total of 608 articles were identified through searching of electronic databases, out of this 554 remained after removing duplications. After screening in title and abstract 112 articles remained; of which 105 articles (review articles and letters to editors) were excluded. Finally, 9 articles were included for meta-analysis^18–26^ (Fig 1). A total number of 3,823 patients were included; 1416 patients on RAAS inhibitors and 3469 on non-RAAS inhibitors (Fig 2). Newcastle-Ottawa Scale methodological quality assessment gave the quality of the included studies in good arrangement (Appendix 2).

**Figure 1:**
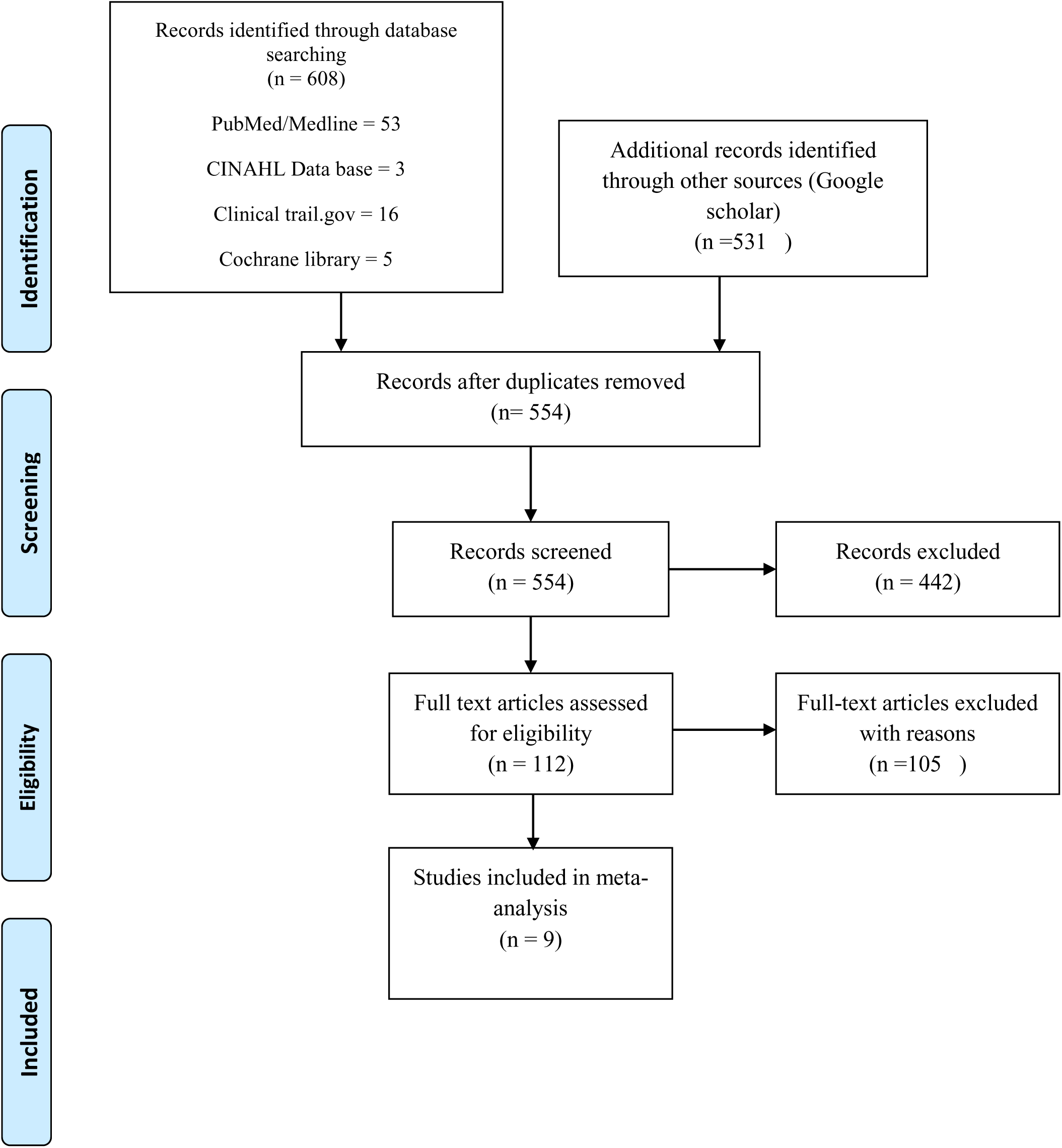
PRISMA flow diagram showing, screened, excluded and included studies All included studies were retrospective, and six were conducted in China, one in USA and one in UK, (table 1).

**Figure 2:**
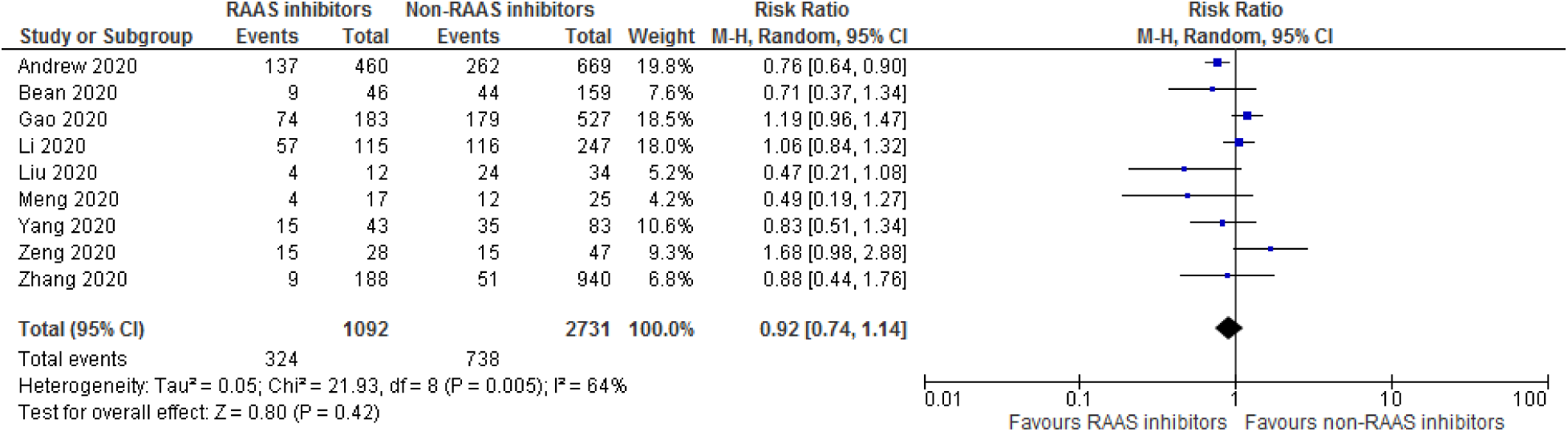
Forest plot for severity of patients taking RAAS inhibitors compared to those not taking RAAS inhibitors.

**Table 1:**
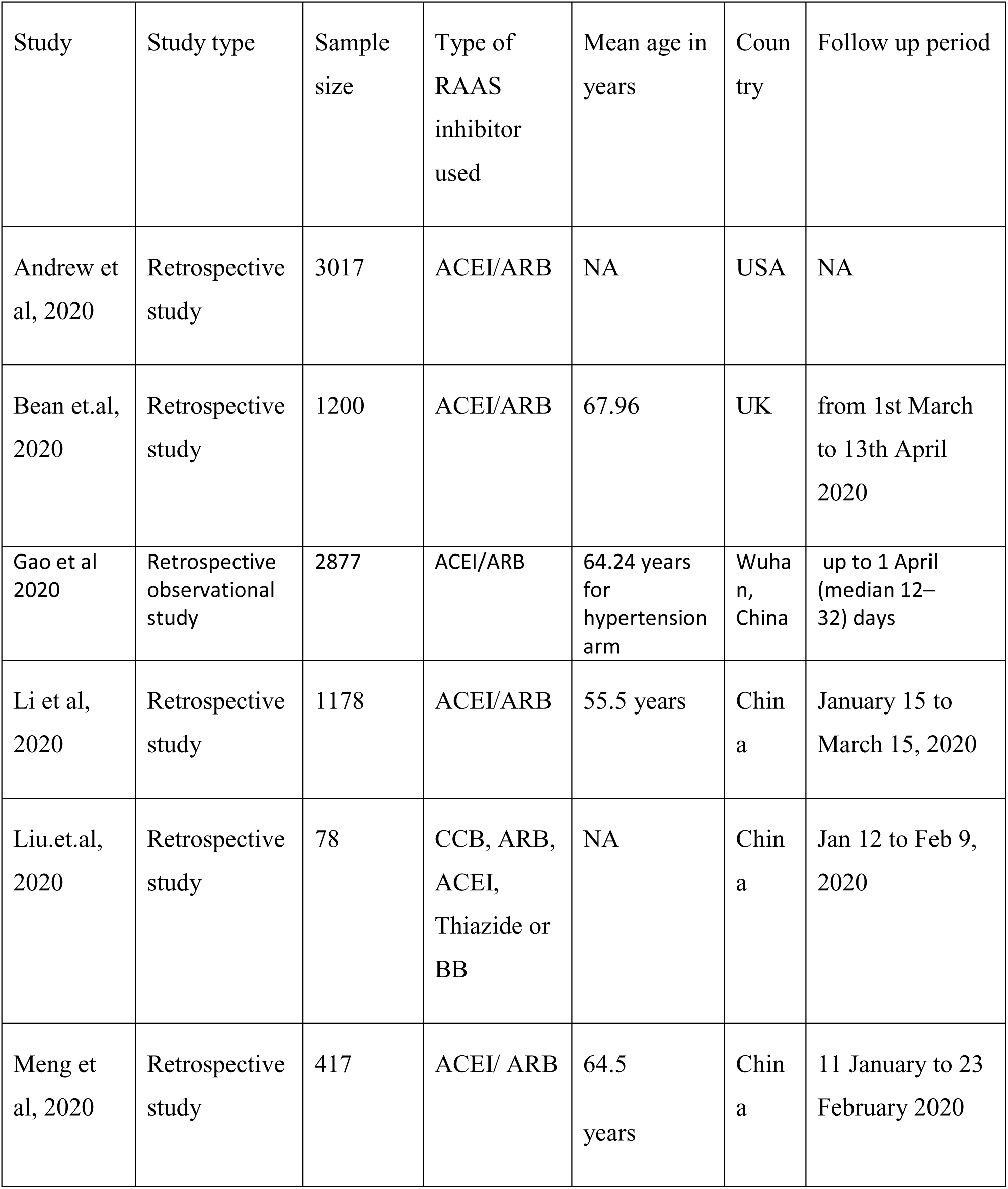
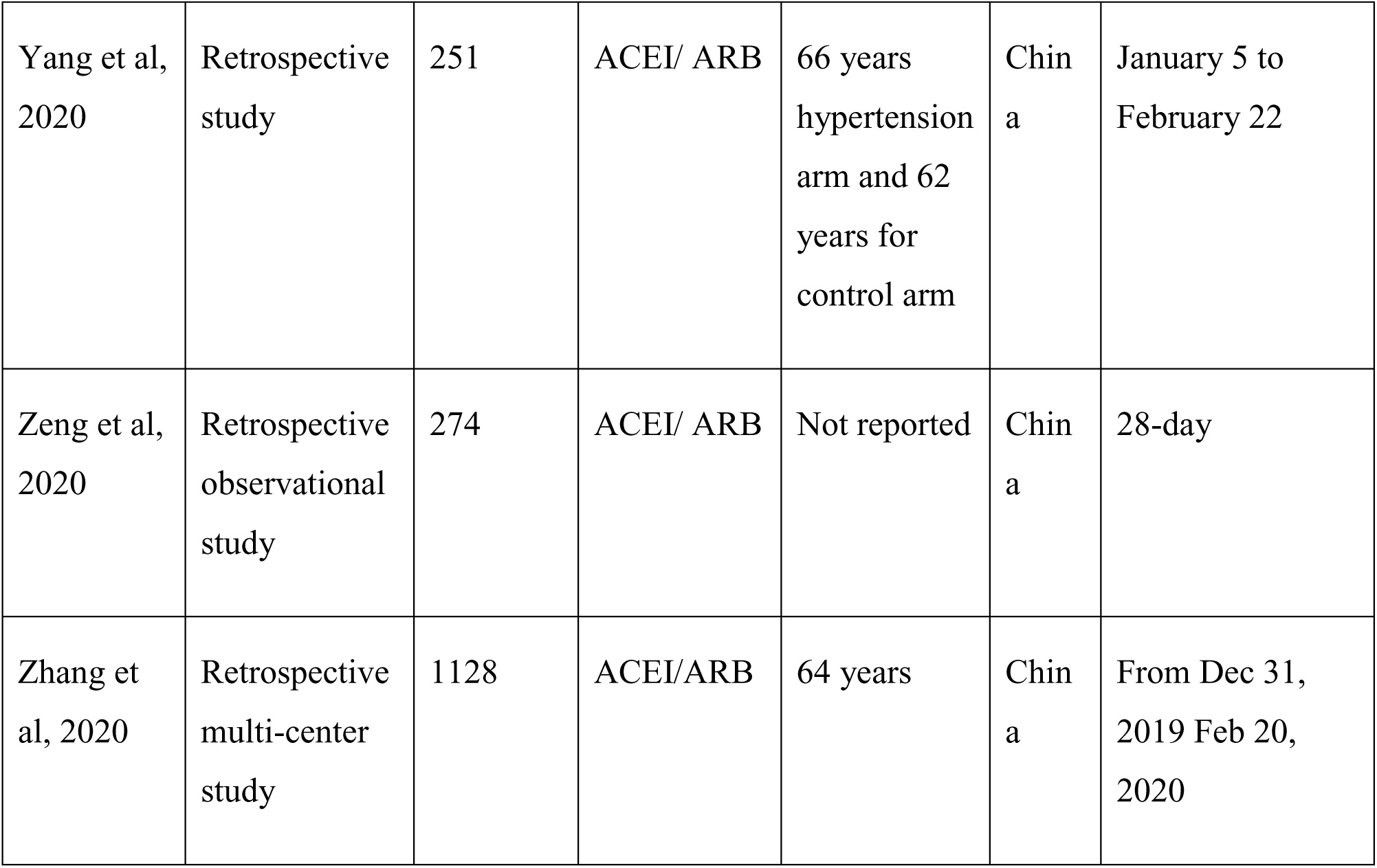
Characteristics of included studies.

| Study | Study type | Sample size | Type of RAAS inhibitor used | Mean age in years | Country | Follow up period |
| --- | --- | --- | --- | --- | --- | --- |
| Andrew et al, 2020 | Retrospective study | 3017 | ACEI/ARB | NA | USA | NA |
| Bean et.al, 2020 | Retrospective study | 1200 | ACEI/ARB | 67.96 | UK | from 1st March to 13th April 2020 |
| Gao et al 2020 | Retrospective observational study | 2877 | ACEI/ARB | 64.24 years for hypertension arm | Wuhan, China | up to 1 April (median 12–32) days |
| Li et al, 2020 | Retrospective study | 1178 | ACEI/ARB | 55.5 years | China | January 15 to March 15, 2020 |
| Liu.et.al, 2020 | Retrospective study | 78 | CCB, ARB, ACEI, Thiazide or BB | NA | China | Jan 12 to Feb 9, 2020 |
| Meng et al, 2020 | Retrospective study | 417 | ACEI/ ARB | 64.5 years | China | 11 January to 23 February 2020 |
| Yang et al, 2020 | Retrospective study | 251 | ACEI/ ARB | 66 years hypertension arm and 62 years for control arm | China | January 5 to February 22 |
| Zeng et al, 2020 | Retrospective observational study | 274 | ACEI/ ARB | Not reported | China | 28-day |
| Zhang et al, 2020 | Retrospective multi-center study | 1128 | ACEI/ARB | 64 years | China | From Dec 31, 2019 Feb 20, 2020 |

### Effect of RAASI on disease severity

Nine studies reported the severity data of the hypertensive COVID-19 patients. No significant association was observed with RAAS inhibitors on severity; RR = 0.92 (95% CI: 0.74–1.14), random effect model. Significant heterogeneity was observed among the studies (I^2^ = = 64%, P = 0.005)Fig. 2.

### Effect of RAAS inhibitors on mortality of patients with COVID_19 and hypertension

Seven studies reported the mortality data of the hypertensive COVID_19 patients. No heterogeneity was observed among the 7 studies (I^2^ = 0%, P = 0.59). The pooled mortality was RR = 0.73 (95% CI: 0.63- 0.85), fixed effect model Fig. 3.

**Figure 3:**
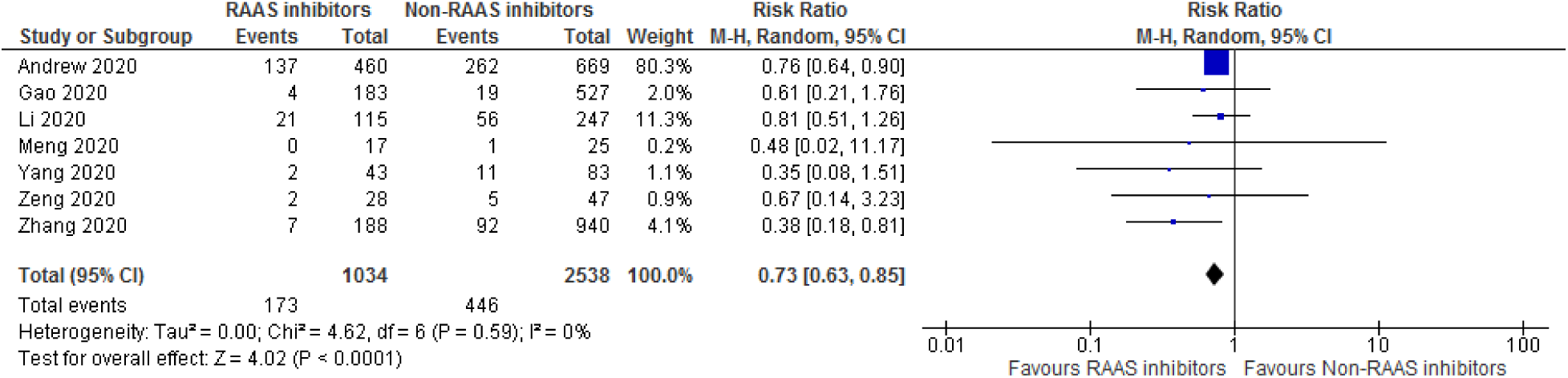
Forest plot for mortality of patients taking RAAS inhibitors compared to not taking RAAS inhibitors

The use of RAAS inhibitor did not have association with long term hospitalization as compared with non RAAS inhibitor users (weighed mean difference –2.33[95% CI: –5.60, 0.75], fixed effect model. No heterogeneity was observed (I^2^ = 0%, P = 0.65), Fig 4.

**Figure 4:**
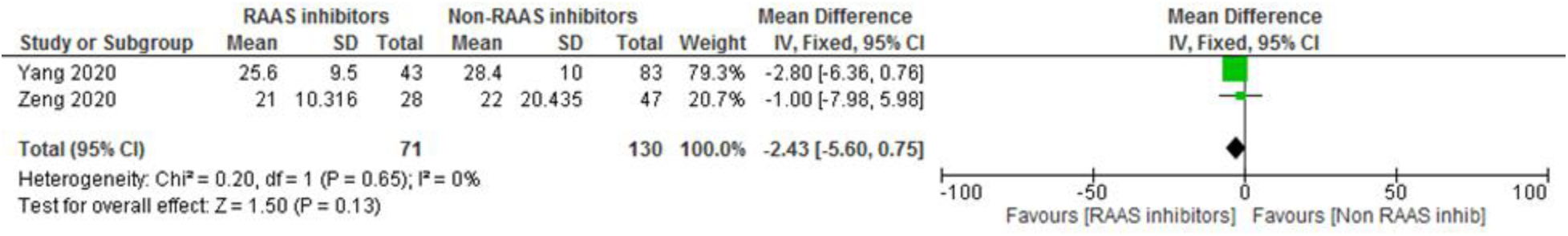
Effect of RAAS inhibitor on the time of hospitalization

### Assessment of bias

The funnel plot was used to assess the publication bias. Funnel plot indicates the existence of small study effects. We performed the regression–based on Egger’s test. The funnel plot was symmetrical in shape for severity and mortality (Fig 5 and 6), indicating no publication bias. Regression-based Egger’s test indicates that there were no small-study effects on mortality (P = 0.122) and severe events (p = 0.686).

**Figure 5:**
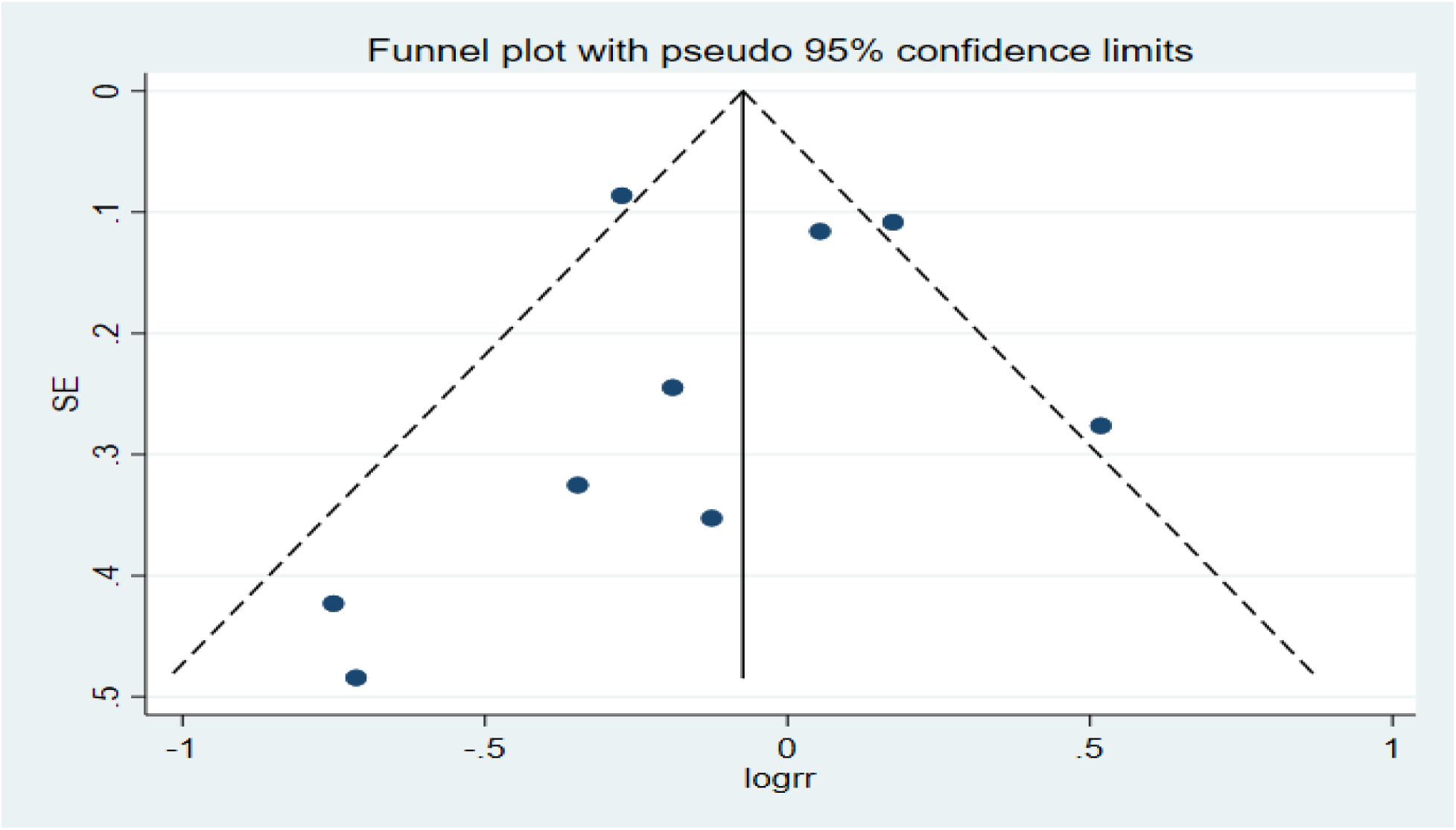
Funnel plot for the severity of diseases

**Figure 6:**
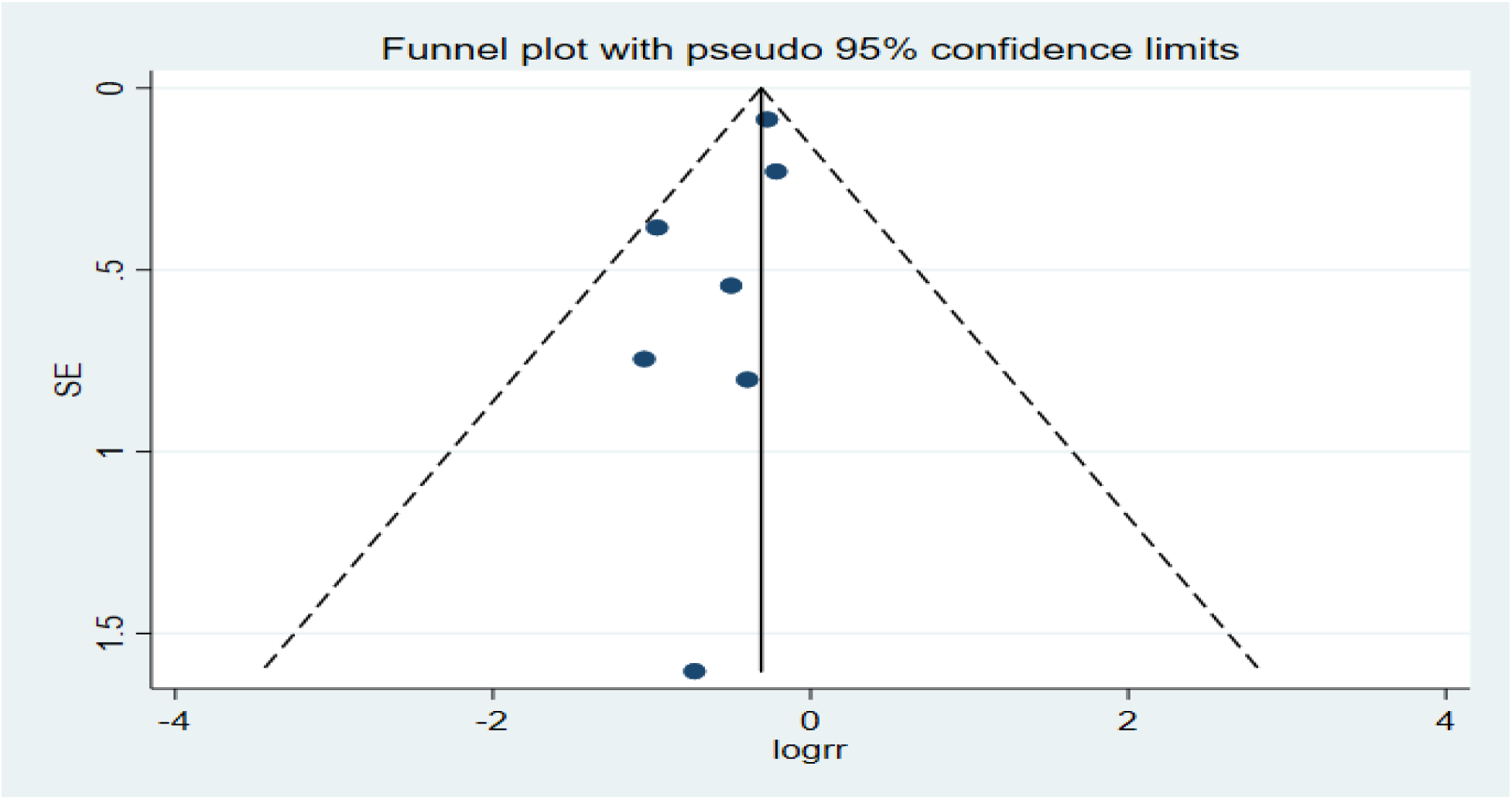
Funnel plot for mortality

## Discussion

Concern over the safety of RAAS inhibitors use in patients contracted COVID_19 originated since SARS-CoV-2 uses ACE2 receptor to enter the target human cell. Animal studies also showed an increase in RAAS inhibitor expression of ACE2 which could speed up the spread of the virus in human cells, predispose for hospitalization, increase and, mortality. This was further strengthened by the report that patients with hypertension taking ACEI/ARB have increased risk of developing severe pneumonia (P = 0.064)^22^. Dooley *et al*. also reported that RAAS inhibitors increased the risk of symptomatic infection with COVID_19 approximately by two-fold in community populations^27^. However, other studies reported that the use of ACEI/ARB did not show association with the outcome of COVID-19 patients^28–31^.

This study, however, was in contrast to the original hypothesis demonstrating the safe usage of RAAS inhibitors in hypertensive COVID_19 patients; in this, it was associated with a significant reduction in mortality. The hypertensive patients with COVID_19 who were on ACE/ARB had a 27% reduction in risk of mortality compared to that not on ACEI/ARB. This finding is helpful and should be considered in clinical decision making. Two large sample size studies,^18^ and^21^, reported a reduction in mortality with patients on ARBs/ACEIs compared to those on non-ARBs/ACEIs medications. A study compared the use of other antihypertensive drugs with ACEI/ARB revealed that ACEI/ARB significantly reduced mortality in COVID_19 patients with hypertension^23^. RAAS inhibitors thought to up-regulate the level of ACE2 which may protect against organ damage by inhibiting the production of angiotensin II^32^. Moreover, the study showed that patients with COVID_19 appeared to exhibit RAAS activation, increased viral load levels, and lung injury^33^. However, all five small sample studies did not show any association with mortality^20–24^. Emerging data suggested that patients with hypertension and diagnosed with COVID_19 are at an increased risk of death^34^. These patients have often prescribed RAAS inhibitors, including ACE inhibitors^35^ and ARBs^36^. The finding of this study indicated the beneficial effect of RAAS inhibitors use over not using them and correlated well with the recommendation of the International Society of Hypertension^37^.

This study showed no significant association in both severities of the disease and hospitalization. Previously reported studies revealed the direct anti-inflammatory effect of RAAS inhibitors which could help in the prevention of cardiovascular complications^38^. RAAS inhibitors reduce pro-inflammatory cytokines, chemokine’s; reduce pro-inflammatory effect leukocyte, and endothelial cells^39^. This could justify the current findings and lower number of critical patients in hypertension patients with COVID-19 on RAAS inhibitors arm. However, fewer study availability and retrospective nature of the included studies warrants further randomized controlled studies and observational studies.

### Conclusion

This study supports the safe use of ACEI/ARB among hypertensive patients contracted COVID_19 and in line with the recommendation of the International Society of Hypertension.

## Data Availability

N/A

## List of abbreviations

COVID_19: Corona viral diseases 2019
SARS-CoV-2: severe acute respiratory syndrome coronavirus 2
CDC: Center of Disease Control
RAAS: renin-angiotensin-aldosterone system
ACEI: Angiotensin-converting enzyme inhibitors
ARB: Angiotensin receptor blocker
ACE2: angiotensin-converting enzyme 2
Ang II: Angiotensin II
PRISMA-P: Systematic Review and Meta-Analysis Protocol
RCTs: randomized controlled trials
CCB: Calcium channel blocker.

## Ethical approval and consent to participate

Not applicable

## Consent for publication

Not applicable

## Authors’ contribution

TBB and TS designed the study, searched literature, analyzed data and drafted manuscript. SD interpreted and reviewed the result. All authors participated, reviewed and approved the final manuscript.

## Acknowledgment

Not applicable

## Appendix 1: Search terms used

(“ace inhibitor*” OR “angiotensin converting enzyme inhibitor*” OR arb*, “angiotensin receptor blocker*” OR captopril OR capoten OR benazepril OR Lotensin OR enalapril OR vasotec OR epaned OR lexxel OR fosinopril OR monopril OR lisinopril OR prinivil OR zestril OR qbrelis OR moexipril OR univasc OR perindopril OR aceon OR quinapril OR accupril OR ramipril OR altace OR trandolapril OR mavik OR teveten OR benicar OR olmesartan OR diovan OR valsartan OR atacand OR candesartan OR cozaar OR losartan OR micardis OR avapro OR edarbi OR irbesartan OR “azilsartan medoxomil” OR prexxartan OR valsartan) AND (“sarscoronavirus” OR “sars-cov-2” OR “severe acute respiratory syndrome” OR coronavir* OR coronovirus* OR “corona virus” OR “corono virus” OR “corono virus” OR “virus corona” OR “virus corono” OR “covid-19” OR “covid19”* OR “covid 19” OR “2019-nCoV” OR (wuhan* AND (virus OR viruses OR viral)) OR (covid* AND (virus OR viruses OR viral)) OR “sars-cov” OR “mers-cov” OR “mers cov” OR “middle east respiratory syndrome”).

## Appendix 2: Newcastle–Ottawa Scale (NOS) methodological quality assessment

| Study | Selection |  |  |  | Comparability | Outcome |  |  | Quality Score |
| --- | --- | --- | --- | --- | --- | --- | --- | --- | --- |
|  | Representativeness of the exposed cohort | Selection of non-exposed cohort | Ascertainment of exposure | Demonstration that outcome of interest was not present at start of study | Comparability of cohorts on the basis of the design or analysis | Assessment of outcome | Was follow up long enough for outcomes to occur | Adequacy of follow up |  |
| Andrew 2020 | * |  | * | * | * | * |  | * | Good |
| Li 2020 | * | * | * |  | * | * | * | * | Good |
| Meng 2020 | * | * | * |  | * | * | * | * | Good |
| Yang 2020 | * | * | * |  | * | * | * | * | Good |
| Zeng 2020 |  | * | * | * | * | * |  | * | Good |
| Zhang 2020 | * | * | * | * | ** | * | * | * | Good |
| Bean 2020 | * | * | * | * | ** | * |  | * | Good |
| Liu 2020 |  | * | * | * | * | * |  | * | Good |
| Gao 2020 | * | * | * | * | ** | * | * | * | Good |

### Interpretation

Good quality: 3 or 4 stars from selection; 1or 2 stars from comparability and 2 or 3 stars from outcome

Fair quality: 2 stars in selection; 1 or 2 stars in comparability and 2 or 3 stars in outcome

Poor quality: 0 or 1star from selection OR 1 or 2 stars in comparability OR 0 or 1 star from outcome

## References

1. Zhu N, Zhang D, Wang W, et al. A novel coronavirus from patients with pneumonia in China, 2019. New England Journal of Medicine. 2020.

2. World Health Organization. Coronavirus disease 2019 (COVID-19): situation report-139. 2020.

3. The Novel Coronavirus Pneumonia Emergency Response Epidemiology Team. The epidemiological characteristics of an outbreak of 2019 novel coronavirus diseases (COVID-19). China2020, p.113–22.

4. Messerli FH, Bangalore S, Bavishi C and Rimoldi SF. Angiotensin-converting enzyme inhibitors in hypertension: to use or not to use? Journal of the American College of Cardiology. 2018; 71: 1474–82.

5. Soler MJ, Barrios C, Oliva R and Batlle D. Pharmacologic modulation of ACE2 expression. Current hypertension reports. 2008; 10: 410.

6. Ferrario CM, Jessup J, Chappell MC, et al. Effect of angiotensin-converting enzyme inhibition and angiotensin II receptor blockers on cardiac angiotensin-converting enzyme 2. Circulation. 2005; 111: 2605–10.

7. Paul M, Poyan Mehr A and Kreutz R. Physiology of local renin-angiotensin systems. Physiological reviews. 2006; 86: 747–803.

8. Hampl V, Herget J, Bíbová J, et al. Intrapulmonary activation of the angiotensin-converting enzyme type 2/angiotensin 1–7/G-protein-coupled Mas receptor axis attenuates pulmonary hypertension in Ren-2 transgenic rats exposed to chronic hypoxia. Physiological research. 2015; 64: 25–38.

9. Li W, Moore MJ, Vasilieva N, et al. Angiotensin-converting enzyme 2 is a functional receptor for the SARS coronavirus. Nature. 2003; 426: 450–4.

10. Zhou P, Yang X-L, Wang X-G, et al. A pneumonia outbreak associated with a new coronavirus of probable bat origin. Nature. 2020; 579: 270–3.

11. Kuba K, Imai Y, Rao S, et al. A crucial role of angiotensin converting enzyme 2 (ACE2) in SARS coronavirus–induced lung injury. Nature medicine. 2005; 11: 875–9.

12. Li W, Zhang C, Sui J, et al. Receptor and viral determinants of SARS‐coronavirus adaptation to human ACE2. The EMBO journal. 2005; 24: 1634–43.

13. Fang L, Karakiulakis G and Roth M. Are patients with hypertension and diabetes mellitus at increased risk for COVID-19 infection? The Lancet Respiratory Medicine. 2020; 8: 21.

14. Diaz JH. Hypothesis: angiotensin-converting enzyme inhibitors and angiotensin receptor blockers may increase the risk of severe COVID-19. Journal of travel medicine. 2020.

15. Sommerstein R, Kochen MM, Messerli FH and Gräni C. Coronavirus Disease 2019 (COVID‐19): Do Angiotensin‐Converting Enzyme Inhibitors/Angiotensin Receptor Blockers Have a Biphasic Effect? Journal of the American Heart Association. 2020; 9: e016509.

16. Ghosal S, Mukherjee JJ, Sinha B and Gangopadhyay KK. The effect of angiotensin converting enzyme inhibitors and angiotensin receptor blockers on death and severity of disease in patients with coronavirus disease 2019 (COVID-19): A meta-analysis. medRxiv. 2020.

17. Liberati A, Altman DG, Tetzlaff J, et al. The PRISMA statement for reporting systematic reviews and meta-analyses of studies that evaluate health care interventions: explanation and elaboration. Journal of clinical epidemiology. 2009; 62: e1–e34.

18. Ip A, Parikh K, Parrillo JE, et al. Hypertension and Renin-Angiotensin-Aldosterone System Inhibitors in Patients with Covid-19. medRxiv. 2020.

19. Li J, Wang X, Chen J, Zhang H and Deng A. Association of Renin-Angiotensin System Inhibitors With Severity or Risk of Death in Patients With Hypertension Hospitalized for Coronavirus Disease 2019 (COVID-19) Infection in Wuhan, China. JAMA cardiology. 2020.

20. Meng J, Xiao G, Zhang J, et al. Renin-angiotensin system inhibitors improve the clinical outcomes of COVID-19 patients with hypertension. Emerging Microbes & Infections. 2020; 9: 757–60.

21. Yang G, Tan Z, Zhou L, et al. Angiotensin II Receptor Blockers and Angiotensin-Converting Enzyme Inhibitors Usage is Associated with Improved Inflammatory Status and Clinical Outcomes in COVID-19 Patients With Hypertension. medRxiv. 2020.

22. Zeng Z, Sha T, Zhang Y, et al. Hypertension in patients hospitalized with COVID-19 in Wuhan, China: A single-center retrospective observational study. medRxiv. 2020.

23. Zhang P, Zhu L, Cai J, et al. Association of inpatient use of angiotensin converting enzyme inhibitors and angiotensin II receptor blockers with mortality among patients with hypertension hospitalized with COVID-19. Circulation Research. 2020.

24. Gao C, Cai Y, Zhang K, et al. Association of hypertension and antihypertensive treatment with COVID-19 mortality: a retrospective observational study. European Heart Journal. 2020; 00: 1–9.

25. Bean D, Kraljevic Z, Searle T, et al. Treatment with ACE-inhibitors is associated with less severe disease with SARS-Covid-19 infection in a multi-site UK acute Hospital Trust. medRxiv. 2020.

26. Liu Y, Huang F, Xu J, et al. Anti-hypertensive Angiotensin II receptor blockers associated to mitigation of disease severity in elderly COVID-19 patients. medRxiv. 2020.

27. Dooley H, Lee K, Freidin M, et al. ACE inhibitors, ARBs and other anti-hypertensive drugs and novel COVID-19: An association study from the COVID Symptom tracker app in 2,215,386 individuals. 2020.

28. Mancia G and Rea F. Renin-Angiotensin-Aldosterone System Blockers and the Risk of Covid-19. 2020.

29. Lee H-Y, Ahn J, Kang CK, et al. Association of Angiotensin II Receptor Blockers and Angiotensin-Converting Enzyme Inhibitors on COVID-19-Related Outcome. Available at SSRN 3569837. 2020.

30. Reynolds HR, Adhikari S, Pulgarin C, et al. Renin–Angiotensin–Aldosterone System Inhibitors and Risk of Covid-19. New England Journal of Medicine. 2020.

31. Tedeschi S, Giannella M, Bartoletti M, et al. Clinical impact of renin-angiotensin system inhibitors on in-hospital mortality of patients with hypertension hospitalized for COVID-19. Clinical Infectious Diseases: An Official Publication of the Infectious Diseases Society of America. 2020.

32. Zou Z, Yan Y, Shu Y, et al. Angiotensin-converting enzyme 2 protects from lethal avian influenza A H5N1 infections. Nature communications. 2014; 5: 3594.

33. Liu Y, Yang Y, Zhang C, et al. Clinical and biochemical indexes from 2019-nCoV infected patients linked to viral loads and lung injury. Science China Life sciences. 2020; 63: 364–74.

34. Zheng Y-Y, Ma Y-T, Zhang J-Y and Xie X. COVID-19 and the cardiovascular system. Nature Reviews Cardiology. 2020; 17: 259–60.

35. von Bayern AMP, Heathcote RJP, Rutz C and Kacelnik A. The role of experience in problem solving and innovative tool use in crows. Curr Biol. 2009; 19: 1965–8.

36. Quinn KL, Fralick M, Zipursky JS and Stall NM. Renin–angiotensin–aldosterone system inhibitors and COVID-19. CMAJ. 2020; 192: E553–E4.

37. International Society of Hypertension. A statement from the International Society of Hypertension on COVID-19. Edinburgh, United Kingdom: International Society of Hypertension, 2020.

38. Montecucco F, Pende A and Mach F. The renin-angiotensin system modulates inflammatory processes in atherosclerosis: evidence from basic research and clinical studies. Mediators of inflammation. 2009; 2009.

39. Dandona P, Dhindsa S, Ghanim H and Chaudhuri A. Angiotensin II and inflammation: the effect of angiotensin-converting enzyme inhibition and angiotensin II receptor blockade. Journal of human hypertension. 2007; 21: 20–7.

